# Mediators of the association between childhood BMI and educational attainment: analysis of a UK prospective cohort study

**DOI:** 10.1101/2022.06.20.22276640

**Authors:** Kirsty Bowman, Tim Cadman, Oliver Robinson, Amanda Hughes, Jon Heron, Alexa Blair Segal, Maria Carmen Huerta, Laura D Howe

**Affiliations:** MRC Integrative Epidemiology Unit at the University of Bristol, Bristol, UK; Population Health Sciences, University of Bristol, Bristol, UK; Department of Public Health, University of Copenhagen, Copenhagen, Denmark; Centre for Health Economics & Policy Innovation, Imperial College Business School, London, UK; MRC Centre for Environment and Health, School of Public Health, Imperial College London, UK

**Keywords:** Children, body mass index, ALSPAC, education attainment, GCSEs, mediation

## Abstract

**Background:** Higher BMI in childhood is associated with lower academic achievement.

**Objective:** To explore potential pathways linking childhood BMI with educational attainment.

**Methods:** In the Avon Longitudinal Study of Parents and Children prospective cohort study (N=6234), we used structural equation models to assess the association between BMI z-scores at 11.7 years and educational attainment at 16 (General Certificate of Secondary Education (GCSE) results). Depressive symptoms, externalising symptoms, bullying, pressure to lose weight, and school enjoyment were considered as potential mediators.

**Results:** Higher BMI z-scores were associated with lower GCSE scores (total effect: females β = -3.20 95% CI -5.15, -1.26; males β = -2.98 95% CI -5.12, -0.84). Depressive symptoms and externalising symptoms partially mediated this association in females (indirect effect: β = -0.35 95% CI -0.63, -0.06, proportion mediated = 11%; β = -0.84 95% CI -1.77, 0.10, proportion mediated = 26%, respectively). In males, there was some evidence that the association was partially mediated by bullying (indirect effect β = -0.35 95% CI -0.74, 0.03, proportion mediated = 12%).

**Conclusions:** The association between childhood BMI and educational attainment may be partially mediated by depressive symptoms and externalising symptoms in females, and by bullying in males.

## Introduction

Globally, the proportion of children and adolescents with obesity has increased dramatically over the past four decades. Between 1975 and 2016 the age standardised prevalence of obesity increased from 0.7 to 5.6% for females and from 0.9 to 7.8% for males aged 2 to 19 years (1). Children with obesity are more likely to suffer from adverse health consequences both in the shorter and longer terms including increased cardiovascular risk factors, cardiovascular disease in later life, and premature mortality (2-4). Moreover, children with obesity are at an elevated risk of transitioning into adulthood with obesity (5).

There is accumulating evidence suggesting that children with obesity are more likely to experience adverse social and economic outcomes. Educational attainment is a particular outcome of interest as it is crucial for employment prospects, later health, wellbeing, and functioning in society (6, 7). Recently, a systematic review focusing on studies using more robust causal inference approaches concluded that childhood obesity hinders educational attainment, with larger negative associations observed for females compared to males (8).

Several pathways have been suggested which may link childhood obesity to poorer educational attainment. Adverse physical consequences of obesity in children may result in feelings of tiredness, thus impairing concentration and productivity, which may affect academic performance (9, 10). Childhood and adolescent obesity has been reported to be associated with attention problems and externalizing behaviours (11, 12), depressive symptoms (especially for females) and major depressive disorder (13-15), and lower self-esteem (16), which could weaken motivation and educational performance (16-19). At the societal level, children and adolescents with obesity are more likely to experience stigmatization from teachers, peers, parents, and the media, and may experience pressure to be thin (20-23). Furthermore, children with obesity may be more likely to have impaired peer relationships, to be bullied, and to report being socially isolated (21, 24, 25). These social factors may reduce both their functioning within their school environment and their academic attainment; for instance, biases from teachers may result in lower teacher-assessed grades (22, 26-28). Children and adolescents with obesity may have impaired school experiences, lower school enjoyment, disengagement and lower participation in learning activities, and lower motivation at school (16, 26, 29). School attendance may also be affected by these consequences of obesity, which in turn may impact academic performance (16).

Elucidating the pathways linking obesity to educational attainment may help to inform strategies to improve the school experience and educational attainment of children with obesity. Overall, there has been a limited number of studies examining potential pathways between childhood obesity and educational attainment (30, 31). Evidence has shown that the following factors may have a mediating role: weight-based teasing (26), behavioural engagement (29), and bullying (9). Furthermore, anxiety, depression and ADHD have been shown to be of importance for educational achievement among children and adolescents with obesity (32). We used data from the Avon Longitudinal Study of Parents and Children, which is an ongoing birth cohort study, to explore potential pathways between childhood body mass index (BMI) and educational attainment. In particular, we sought to understand the mediating roles of depressive symptoms, bullying, externalising behaviours (hyperactivity and conduct problems), pressure to lose weight and school enjoyment, and we explored the sex differences in these pathways.

## Methods

### Study participants

This analysis used data from the Avon Longitudinal Study of Parents and Children (ALSPAC) which is a birth cohort within the South West of England. Pregnant women were invited to take part in the study if they were resident in the former Avon area with an estimated delivery date between 1st April 1991 and 31st December 1992. The core sample comprises of 14,541 pregnant women with 13,988 children alive at 12 months. There have been regular follow-ups via questionnaires and clinical assessment of the children and parents since the initiation of the cohort. Further detailed descriptions of the cohort are given elsewhere (33, 34). The study website contains details of all the data which is available through a fully searchable data dictionary and variable search tool (http://www.bristol.ac.uk/alspac/researchers/our-data/). Ethics approval for the study was obtained from the ALSPAC Ethics and Law Committee and the local research ethics committees. At age 18, study children were sent ‘fair processing’ materials describing ALSPAC’s intended use of their health and administrative records and were given clear means to consent or object via a written form. Data were not extracted for participants who objected, or who were not sent fair processing materials.

### Exposure: Body mass index

Children attended a clinical assessment centre at mean age 11.7 years. Height was measured, where possible without shoes, using a Harpenden Stadiometer and recorded to the last complete millimetre. Weight was measured, where possible in underclothes and without shoes, using a Tanita Body Fat Analyser (Model TBF 305) to the nearest 50 grammes. The BMI measures, calculated as weight in kilogrammes divided by height in metres squared, were age- and sex-standardized to the 1990 UK Growth Reference.

### Potential mediators

#### Depressive symptoms

Children attended a clinical assessment centre at mean age 13.8 years. Depressive symptoms were assessed via self-report using the short version of the Mood and Feelings Questionnaire (SMFQ) (35). The questionnaire has thirteen statements asking about symptoms experienced in the previous two weeks with the following choice of response options: “true”, “sometimes”, or “not at all” (Tables S1 and S2). The responses for the 13 statements in this analysis were coded so that higher scores reflect greater depressive symptoms.

#### Externalising symptoms

Externalising symptoms were based on the mother-rated hyperactivity and conduct subscales of the Strengths and Difficulties (SDQ) questionnaire (36) which was completed when the study children were mean age 13.2 years. Each of the two subscales consist of five statements relating to children’s difficulties within the previous six months with the following response options: “not true”, “somewhat true”, or “certainly true” (Tables S1 and S2). Responses for the ten statements in this analysis were coded so that higher scores reflect greater externalising symptoms.

#### School enjoyment

Children completed a questionnaire at mean age 14.2 years. The questionnaire had several items capturing school enjoyment e.g. (how much the respondent’s school is a place where they enjoy what they do in class). The questions centred around the child’s enjoyment of classes and school with the response options: “strongly agree”, “agree”, “disagree”, “strongly disagree” (Tables S1 and S2). The responses for these three statements were coded so that higher scores reflect lower school enjoyment.

#### Bullying

Children attended a clinical assessment centre at mean age 12.8 years. Bullying was assessed using a modified version of the Bullying and Friendship Interview Schedule (BFIS). This analysis uses the sections on received overt (direct) bullying to establish victimisation and received relational (indirect) bullying to establish relational victimisation. In this analysis we used four of the statements in the received overt bullying and four statements of the received relational bullying (Tables S1 and S2). We did not include the following experiences in our analysis due to small numbers of participants reporting these events: “Someone has done other bad things to teenager”, “Friends have done other things to upset teenager”, and “Frequency someone has hit/beaten up teenager: friends and peers”. Children were asked if they had experienced any of the events in the last six months and if so, follow-up questions were asked about the frequency. Response options for the frequency were: “seldom (1 to 3 times)”, “frequently (> 4 times)”, and “very frequently (>1/week)”. Children who responded that they had not experienced that event were coded as a frequency of never. In this analysis we combined both received overt and received relational bullying and the responses were coded so that higher scores reflect greater peer victimisation.

#### Pressure to lose weight

Children completed a questionnaire at mean age 13.9 years. Pressure to lose weight was established from four questionnaire items relating to pressure to lose weight from friends, family, partners, and the media. The response options for these statements were: “not at all”, “yes, a little”, “yes, quite a lot”, and “yes, a lot” (Tables S1 and S2). The responses to these four statements were coded in this analysis so that higher scores reflect greater pressure to lose weight.

#### Outcome: Educational attainment

Educational attainment was assessed using a summary measure based on General Certificate of Secondary Education (GCSE) qualifications taken at age 16, which represented the end of compulsory education for study participants. Results from GCSEs and equivalent qualifications were obtained from linkage between ALSPAC and the National Pupil Database. The summary measure, a total GCSE and equivalents ‘capped’ score, is calculated by converting qualification grades into points (e.g. an A* in one subject is worth 58 points), and calculating the total from the student’s best eight subjects. This is to prevent scores being inflated by students who sit more qualifications. The possible range from 8 GCSEs is 0 to 464, but a small number of students who took more advanced qualifications early had scores between 464 and 540.

#### Covariates

The following variables were considered as possible confounders of the association between the child’s BMI and their educational attainment: maternal age at pregnancy (years), maternal smoking in pregnancy (categorised as a binary variable, yes/no), housing tenure (categorised as a binary variable, mortgaged/owned/rented privately/other vs council rented/housing association rented), highest maternal education qualification (none/Certificate of Secondary Education (CSE)/vocational; O-level; A-level/degree/higher), maternal occupational social class (professional, managerial and technical; skilled non-manual/skilled manual; partly skilled/unskilled) and parity (0 vs ≥1). Maternal smoking, parity, social class, housing tenure and highest education were derived from questionnaires completed during the antenatal period.

### Statistical analyses

We restricted our analysis to participants who had height and weight measured at a minimum of one research clinic visit between age 7 and age 15 (six potential measurement occasions, with measures other than the main exposure at age 11.7 years used as auxiliary variables in multiple imputation analyses) and those who had a GCSE and equivalents capped score in the National Pupil Database (For further detail see Missing data section and for further details on the imputation model see Supporting information (Tables S1 and S2)). We excluded participants who had a record documented in Key Stage 4 of having either Special Education Need (SEN) School Action or a SEN Action Plus (n = 1,320), as we hypothesised associations may differ in these children. We further excluded participants with a missing a record of ethnic background (n = 1,658) because we did not have sufficient auxiliary data to impute ethnicity. Our analysis therefore included 6,234 participants (Figure S1).

Analyses were conducted using STATA version 15 and R version 3.61. Structural equation modelling (SEM) was used to estimate the relationships between childhood BMI z-scores and GCSE and equivalents capped score (R ‘lavaan package’). Potential mediators - depressive symptoms, externalising symptoms, bullying, pressure to lose weight, and school experience - were all modelled as latent variables, with each questionnaire item relating to a particular mediator as a separate indicator variable for that latent variable (Figure 1). The potential mediators were examined individually. The proportion mediated reflects the indirect effect divided by the total effect. Model fit was assessed using the Root Mean Square Error of Approximation (RMSEA), the comparative Fit Index (CFI), and Tucker-Lewis Index (TLI). All analyses were run separately for males and females due to prior evidence that associations of BMI with educational attainment and some of the mediators may differ by sex (8, 37). As a sensitivity analysis, we restricted analysis to participants of white ethnicity.

**Figure 1.** Example of structural equation model used to assess mediation. A simplified diagram illustrating the structural equation models used in analyses. The mediator (here, pressure to lose weight) is modelled as a latent (unobserved) variable, with each relevant questionnaire item informing the latent variable. The curved arrow from BMI to educational attainment reflects the ‘direct effect’; the two straight arrows from BMI to pressure to lose weight and from pressure to lose weight to educational attainment reflect the indirect effect.

### Missing data

To deal with missing data on the exposure, the mediators or the covariates, we used multivariate multiple imputation. Prior to imputation we combined categories where there were low cell counts (n <50). Multiple imputation was used to create data sets for the females and males separately (m = 20). We used multiple chained equations (mice) to impute missing data for all the variables. The imputation model contained all the variables to be included in the SEM models and variables known to be predictors of missingness. For further details on the imputation model see Supporting information material (Tables S1 and S2). Estimates from the SEM models were combined using Rubin’s rules (38).

## Results

### Descriptive statistics

Table 1 shows the characteristics of the imputed sample. The mean BMI z-score (using the 1990 UK reference) at age 11.7 years was 0.32 (SD 1.22) for the females and 0.41 (SD 1.21) for the males. The mean GCSE point score was 353.2 (SD 73.09) for the females and 337.9 (SD 79.02) for the males (Table 1). The distributions of the variables in the imputed sample tended to be similar to those observed in the raw data (Tables S1 and S2).

**Table 1:** Characteristics of participants included in analysis; females (n = 3,308) and males (n = 2,926) using imputed data

|  | Females | Males |
| --- | --- | --- |
| BMI z-score at 11.7 years (mean (SD)) | 0.32 (1.22) | 0.41 (1.21) |
| Maternal smoking in pregnancy (%) | 18.6 | 18.6 |
| Maternal social class (%) |  |  |
| (I – professional) | 3.1 | 3.8 |
| (II - Managerial and technical) | 30.7 | 30.7 |
| (IIINM - Skilled non-manual) | 43.2 | 44.3 |
| (IIIM - Skilled manual) | 3.5 | 3.8 |
| (IV - Partly skilled) | 15.8 | 14.5 |
| (V – Unskilled) | 3.7 | 2.9 |
| Maternal education (%) |  |  |
| (Degree) | 13.6 | 14.0 |
| (A-level) | 25.7 | 26.0 |
| (O-level) | 36.5 | 36.9 |
| (vocational) | 9.3 | 9.1 |
| (none/CSE) | 14.9 | 14.0 |
| Maternal housing tenure (Council rented/ Housing association rented) (%) | 10.1 | 9.0 |
| Parity (≥1) (%) | 53.9 | 53.5 |
| Maternal age at birth (mean (SD)) | 28.6 (4.53) | 29.0 (4.59) |
| Capped GCSE point score (mean (SD)) | 353.2 (73.09) | 337.9 (79.02) |

### Model fit for SEMs

The fit of the latent variable models was moderate to good; taking average fit statistics across the 20 imputed datasets, model fit in females when the mediator was externalising symptoms: RMSEA = 0.08, CFI = 0.93, TLI = 0.91; model fit in males when the mediator was bullying: RMSEA = 0.01, CFI = 1.00, TLI = 1.00 (Table S3). The factor loadings for all the items contributing to the latent variables for mediators were moderate to high (females: 0.60 – 1.37 and males: 0.64 – 1.42) (Table S4).

### Associations of BMI with potential mediators, and of potential mediators with educational attainment

Table 2 shows the association of the BMI z-scores with each of the potential mediators, and the associations of each of the potential mediators with GCSE scores. For females, higher BMI z-scores were associated with depressive symptoms (β = 0.06 95% CI 0.03 to 0.09), and greater pressure to lose weight (β = 0.37 95% CI 0.33 to 0.41). There was also weak evidence of an association between higher BMI z-score and higher externalising symptoms (β = 0.02 95% CI -0.00 to 0.04). In males, higher BMI was associated with higher pressure to lose weight (β = 0.49 95% CI 0.42 to 0.56) and higher levels of bullying (β = 0.04 95% CI 0.01 to 0.07).

**Table 2:** Associations of BMI with potential mediators, and potential mediators with educational attainment. Pooled results for the females (n = 3,308) and males (n = 2,926) from structural equation models (SEM)

|  | Association of BMI with mediator |  |  | Association of mediator with GCSE score |  |  |
| --- | --- | --- | --- | --- | --- | --- |
| | $\beta^1$ | 95% CI | p-value | $\beta^1$ | 95% CI | p-value |
| **Females** |  |  |  |  |  |  |
| Depressive Symptoms | 0.06 | ( 0.03 to 0.09 ) | <0.001 | -6.08 | ( -10.12 to -2.04 ) | 0.003 |
| Lack of school enjoyment | -0.00 | ( -0.04 to 0.03 ) | 0.9 | -24.43 | ( -28.52 to -20.34 ) | <0.001 |
| Pressure to lose weight | 0.37 | ( 0.33 to 0.41 ) | <0.001 | 0.86 | ( -3.66 to 5.38 ) | 0.7 |
| Externalising symptoms | 0.02 | ( -0.00 to 0.04 ) | 0.08 | -40.48 | ( -46.22 to -34.73 ) | <0.001 |
| Bullying | 0.01 | ( -0.02 to 0.04 ) | 0.6 | -7.04 | ( -12.46 to -1.62 ) | 0.01 |
| **Males** |  |  |  |  |  |  |
| Depressive Symptoms | 0.01 | ( -0.01 to 0.04 ) | 0.3 | -19.56 | ( -25.05 to -14.07 ) | <0.001 |
| Lack of school enjoyment | -0.00 | ( -0.04 to 0.04 ) | 0.9 | -12.24 | ( -18.14 to -6.33 ) | <0.001 |
| Pressure to lose weight | 0.49 | ( 0.42 to 0.56 ) | <0.001 | -2.76 | ( -8.47 to 2.94 ) | 0.3 |
| Externalising symptoms | 0.01 | ( -0.01 to 0.03 ) | 0.4 | -48.27 | ( -55.34 to -41.21 ) | <0.001 |
| Bullying | 0.04 | ( 0.01 to 0.07 ) | 0.006 | -8.67 | ( -15.85 to -1.49 ) | 0.02 |

<sup>1</sup> unstandardized linear regression coefficients. Models were adjusted for maternal age at pregnancy (years), maternal smoking in pregnancy, housing tenure, highest maternal education qualification, maternal social class, and parity. Potential mediators are all continuous latent variables.

Higher levels of depressive symptoms, lower school enjoyment, higher externalising symptoms and bullying were all associated with lower educational attainment in both males and females (Table 2).

### Mediation of the association between BMI and educational attainment

Higher BMI z-scores at age 11.7 years were associated with lower GCSE scores (total effect: females β = -3.20 95% CI -5.15 to -1.26 and males β = -2.98 95% CI -5.12 to -0.84) (Table 3). When each mediator was considered separately, there was evidence that this association was partly mediated by depressive symptoms and externalising symptoms in females (indirect effect for depressive symptoms: β = -0.35 95% CI -0.63 to -0.06, proportion mediated 0.11; indirect effect for externalising symptoms: β = -0.84 95% CI -1.77 to 0.10, proportion mediated 0.26) (Table 3). In males, there was some evidence that the association between higher BMI and lower education was partially mediated by bullying (indirect effect β = -0.35 95% CI -0.74 to 0.03, proportion mediated 0.12). The indirect effects for all other mediators were imprecisely estimated, with wide confidence intervals.

**Table 3:** Association between BMI z-score and educational attainment (GCSE point score), and mediation of this association; females (n = 3,308) and males (n = 2,926). Each mediator is considered separately in a structural equation model

| **Females** |  |  |  |  |  |  |  |
| --- | --- | --- | --- | --- | --- | --- | --- |
| *Overall association between BMI z-score and educational attainment (total effect)* |  |  |  |  |  |  |  |
| | $\beta^1$ | 95% CI | p-value | | | | |
|  | -3.20 | ( -5.15 to -1.26 ) | 0.001 |  |  |  |  |
| *Mediation of the association between BMI z-score and educational attainment* |  |  |  |  |  |  |  |
|  | *Direct effect (association between BMI and educational attainment after adjusting for the mediator)* |  |  | *Indirect effect (association between BMI and educational attainment that operates through the mediator)* |  |  | *Proportion mediated<sup>2</sup>* |
| | $\beta^1$ | 95% CI | p-value | $\beta^1$ | 95% CI | p-value | |
| Depressive Symptoms | -2.85 | ( -4.80 to -0.91 ) | 0.004 | -0.35 | ( -0.63 to -0.06 ) | 0.02 | 0.11 |
| Lack of school enjoyment | -3.24 | ( -5.14 to -1.34 ) | <0.001 | 0.04 | ( -0.79 to 0.87 ) | 0.9 | NA |
| Pressure to lose weight | -3.52 | ( -6.08 to -0.95 ) | 0.007 | 0.32 | ( -1.35 to 1.99 ) | 0.7 | NA |
| Externalising symptoms | -2.36 | ( -4.25 to -0.48 ) | 0.01 | -0.84 | ( -1.77 to 0.10 ) | 0.08 | 0.26 |
| Bullying | -3.14 | ( -5.10 to -1.19 ) | 0.002 | -0.06 | ( -0.26 to 0.15 ) | 0.6 | 0.02 |
| **Males** |  |  |  |  |  |  |  |
| *Overall association between BMI z-score and educational association (total effect)* |  |  |  |  |  |  |  |
| | $\beta^1$ | 95% CI | p-value | | | | |
|  | -2.98 | ( -5.12 to -0.84 ) | 0.006 |  |  |  |  |
|  | *Direct effect (association between BMI and educational attainment after adjusting for the mediator)* |  |  | *Indirect effect (association between BMI and educational attainment that operates through the mediator)* |  |  | *Proportion mediated<sup>2</sup>* |
| | $\beta^1$ | 95% CI | p-value | $\beta^1$ | 95% CI | p-value | |
| Depressive Symptoms | -2.69 | ( -4.80 to -0.57 ) | 0.01 | -0.29 | ( -0.86 to 0.28 ) | 0.3 | 0.10 |
| Lack of school enjoyment | -2.99 | ( -5.15 to -0.84 ) | 0.006 | 0.01 | ( -0.53 to 0.56 ) | 0.9 | NA |
| Pressure to lose weight | -1.61 | ( -5.14 to 1.92 ) | 0.4 | -1.37 | ( -4.20 to 1.46 ) | 0.3 | 0.46 |
| Externalising symptoms | -2.55 | ( -4.61 to -0.49 ) | 0.02 | -0.43 | ( -1.53 to 0.67 ) | 0.4 | 0.14 |
| Bullying | -2.63 | ( -4.77 to -0.49 ) | 0.02 | -0.35 | ( -0.74 to 0.03 ) | 0.07 | 0.12 |

<sup>1</sup>mean difference in GCSE point score. Models were adjusted for maternal age at pregnancy (years), maternal smoking in pregnancy, housing tenure, highest maternal education qualification, maternal social class, and parity. The mediators are all continuous latent variables.
<sup>2</sup>the proportion mediated is not calculated where there is inconsistent mediation, i.e. where the indirect effect is positive and the total effect is negative

In a sensitivity analysis restricting the analysis to those of white British ethnicity, the results showed a similar pattern to the main analysis (Table S5).

## Discussion

In this cohort study, each one SD higher BMI was associated with a reduction in the GCSE score of approximately 3 points. This represents about 0.04 standard deviations of the score, so the magnitude of this association is small. In females, approximately 11% of the association between BMI and educational attainment was mediated by depressive symptoms, and 26% by externalising symptoms. In males, approximately 12% of the association between BMI and educational attainment was mediated by bullying. The stigma associated with obesity is likely to contribute to associations between higher BMI and depression, externalising symptoms and bullying (39, 40). In turn, depression, externalising symptoms, and bullying may influence educational attainment through a set of inter-linked mechanisms, including school connectedness, social support, engagement with learning, and attendance (41). Efforts to address weight-related stigma, as well as support for mental health and anti-bullying initiatives may therefore help to alleviate the adverse social, mental health, and educational consequences experienced by children and young people with obesity. The majority of the association between BMI and educational attainment was not explained by the mediators we considered, suggesting other pathways are important. Self-esteem, a key potential mediator, could not be explored due to a lack of data at the appropriate time points.

Previous analyses exploring potential pathways between BMI and educational attainment have also suggested that weight-based teasing (26) and bullying (9) may have a mediating role. Here we found some evidence of a role for bullying in males, but not females, although educational attainment was assessed at an earlier age in the previous studies (9.5 years and 12 years respectively, as opposed to 16 years in the current study). A former analysis, also using ALSPAC, examined individual mediators in relation to BMI and educational attainment, and found there was no mediation via depression for females and males (42). Depressive symptoms were assessed at age 11, which is earlier than the current study, and before the age at which depressive symptoms typically begin to rise most rapidly (43).

The sex differences in the roles of mediators are of interest. Depression was associated with lower educational attainment in both males and females, with a stronger association in males. However, BMI was associated with depression only in females, consistent with other evidence showing stronger effects of BMI on depression in females compared with males (37). Similarly, externalising symptoms was associated with lower educational attainment in both females and males, but the association between BMI and externalising symptoms was stronger in females compared with males. In contrast, bullying showed similar associations with educational attainment in females and males, but the association between BMI and bullying was stronger in males. This finding contrasts with a meta-analysis that found overweight and obese boys and girls were equally likely to experience bullying (25).

Analyses using genetic data as instrumental variables (Mendelian randomization) have suggested that the association between BMI and educational attainment may be causal (44-46). However, recent studies that have applied Mendelian randomization approaches to family-level data have suggested that biases due to family-level processes may drive this association. These processes may include intergenerational effects, including influence of parental BMI on children’s outcomes, and non-random partnership in the parent’s generation with respect to educational attainment or body weight (assortative mating) (47). It is therefore possible that the associations observed in this study in part reflect family-level processes, and not only the influence of a child’s BMI.

### Strengths and limitations

We used data for over 6,000 children from a large birth cohort. The large sample size enabled analysis of males and females separately, given our a priori hypothesis that both the association between BMI and educational attainment and the mediating pathways may differ by sex. A strength of this analysis was using objective research clinic-based measures of BMI and linked educational data, thereby minimising measurement error in the exposure and outcome. Measurement error in a mediator can lead to an underestimation of the indirect effect (48). Mediators were self-or mother-reports from questionnaires, but we used structural equation modelling with latent variables defined by multiple indicator variables for each mediator, helping to reduce the influence of measurement error on our analyses. We attempted to control for known confounders. However, as this was an observational study, there may still be residual confounding, including confounders of the mediator-outcome association, which can induce collider bias (49). For some of the mediators there was a high proportion of missing data, and thus we used multiple imputation to maximise statistical power and reduce selection bias. ALSPAC is a birth cohort study of those who were resident in the former Avon area (South West England) and thus is not nationally representative; there are higher proportions of those from higher socioeconomic status and white British ethnic groups, which could limit the generalizability of these results (33, 50).

## Conclusions

Higher BMI z-scores at age 11.7 years were associated with lower educational attainment. The association between childhood BMI and educational attainment may be partially mediated by depressive symptoms and externalising symptoms in females, and by bullying in males.

## Data Availability

The data concerns individual human subjects and therefore is not freely available, but can be obtained by researchers by applying to ALSPAC.

## Acknowledgements

Kirsty Bowman, Tim Cadman, Jon Heron, Maria Carmen Huerta, and Laura D Howe were involved in the study concept and design. Kirsty Bowman, Tim Cadman, and Amanda Hughes were involved in the data preparation. Kirsty Bowman analysed the data. Kirsty Bowman and Laura D Howe drafted the manuscript. All authors were involved in interpretation of the data, critical revision of the manuscript, and read and approved the final manuscript. This research was funded by the Health Foundation as part of their Social and Economic Value of Health programme. The Health Foundation is an independent charity committed to bringing about better health and health care for people in the UK. Laura D Howe is supported by a Career Development Award from the UK Medical Research Council (MR/M020894/1). Tim Cadman received funding from the European Union’s Horizon 2020 research and innovation programme under grant agreement No 733206, LIFE-CYCLE project. The UK Medical Research Council and Wellcome Trust (Grant ref: 102215/2/13/2) and the University of Bristol provide core support for ALSPAC. A comprehensive list of grants funding is available on the ALSPAC website (http://www.bristol.ac.uk/alspac/external/documents/grant-acknowledgements.pdf).

We are extremely grateful to all the families who took part in this study, the midwives for their help in recruiting them, and the whole ALSPAC team, which includes interviewers, computer and laboratory technicians, clerical workers, research scientists, volunteers, managers, receptionists and nurses.

## References

1. NCD Risk Factor Collaboration. Worldwide trends in body-mass index, underweight, overweight, and obesity from 1975 to 2016: a pooled analysis of 2416 population-based measurement studies in 128·9 million children, adolescents, and adults. Lancet. 2017; 390: 2627–2642.

2. Lawlor DA, Benfield L, Logue J, et al. Association between general and central adiposity in childhood, and change in these, with cardiovascular risk factors in adolescence: prospective cohort study. BMJ. 2010; 341: c6224.

3. Baker JL, Olsen LW, Sørensen TIA. Childhood Body-Mass Index and the Risk of Coronary Heart Disease in Adulthood. N Engl J Med. 2007; 357: 2329–2337.

4. Park MH, Falconer C, Viner RM, Kinra S. The impact of childhood obesity on morbidity and mortality in adulthood: a systematic review. Obes Rev. 2012; 13: 985–1000.

5. Simmonds M, Llewellyn A, Owen CG, Woolacott N. Predicting adult obesity from childhood obesity: a systematic review and meta-analysis. Obes Rev. 2016; 17: 95–107.

6. Easterbrook MJ, Kuppens T, Manstead ASR. The Education Effect: Higher Educational Qualifications are Robustly Associated with Beneficial Personal and Socio-political Outcomes. Social Indicators Research. 2015; 126: 1261–1298.

7. Ross CE, Wu C. The Links Between Education and Health. Am Sociol Rev. 1995; 60(5): 719–745.

8. Segal AB, Huerta MC, Aurino E, Sassi F. The impact of childhood obesity on human capital in high-income countries: A systematic review. Obes Rev. 2021; 22: 1–15.

9. Veldwijk J, Fries MCE, Bemelmans WJE, et al. Overweight and school performance among primary school children: the PIAMA birth cohort study. Obesity. 2012; 20(3): 590–596.

10. Black N, Johnston DW, Peeters A. Childhood Obesity and Cognitive Achievement. Health Econ. 2015; 24: 1082–1100.

11. Pulgarón ER. Childhood obesity: a review of increased risk for physical and psychological comorbidities. Clin Ther. 2013; 35(1): A18–32.

12. Halfon N, Larson K, Slusser W. Associations between obesity and comorbid mental health, developmental, and physical health conditions in a nationally representative sample of US children aged 10 to 17. Acad Pediatr. 2013; 13(1): 6–13.

13. Rao WW, Zong QQ, Zhang JW, et al. Obesity increases the risk of depression in children and adolescents: Results from a systematic review and meta-analysis. J Affect Disord. 2020; 267: 78–85.

14. Quek YH, Tam WWS, Zhang MWB, Ho RCM. Exploring the association between childhood and adolescent obesity and depression: a meta-analysis. Obes Rev. 2017; 18: 742–754.

15. Sutaria S, Devakumar D, Yasuda SS, Das S, Saxena S. Is obesity associated with depression in children? Systematic review and meta-analysis. Arch Dis Child. 2019; 104: 64–74.

16. An R, Yan H, Shi X, Yang Y. Childhood obesity and school absenteeism: a systematic review and meta-analysis. Obes Rev. 2017; 18: 1412–1424.

17. Livaditis M, Zaphiriadis K, Samakouri M, Tellidou C, Tzavaras N, Xenitidis K. Gender Differences, Family and Psychological Factors Affecting School Performance in Greek Secondary School Students. Educ Psychol. 2003; 23(2): 223–231.

18. Sigfúsdóttir ID, Kristjánsson AL, Allegrante JP. Health behaviour and academic achievement in Icelandic school children. Health Educ Res. 2007; 22(1): 70–80.

19. Lundy SM, Silva GE, Kaemingk KL, Goodwin JL, Quan SF. Cognitive Functioning and Academic Performance in Elementary School Children with Anxious/Depressed and Withdrawn Symptoms. Open Pediatr Med Journal. 2010; 4: 1–9.

20. McCabe MP, Ricciardelli LA. A prospective study of pressures from parents, peers, and the media on extreme weight change behaviors among adolescent boys and girls. Behav Res Ther. 2005; 43: 653–668.

21. Rees RW, Caird J, Dickson K, Vigurs C, Thomas J. ‘It’s on your conscience all the time’: a systematic review of qualitative studies examining views on obesity among young people aged 12-18 years in the UK. BMJ Open. 2014; 4: e004404.

22. Puhl RM, Heuer CA. The stigma of obesity: a review and update. Obesity. 2009; 17(5): 941–964.

23. Nutter S, Ireland A, Alberga AS, et al. Weight Bias in Educational Settings: a Systematic Review. Curr Obes Rep. 2019; 8: 185–200.

24. Janssen I, Craig WM, Boyce WF, Pickett W. Associations Between Overweight and Obesity with Bullying Behaviours in School-Aged Children. Pediatrics. 2004; 113(5): 1187–1194.

25. van Geel M, Vedder P, Tanilon J. Are overweight and obese youths more often bullied by their peers? A meta-analysis on the correlation between weight status and bullying. Int J Obes. 2014; 38: 1263–1267.

26. Krukowski RA, West DS, Philyaw Perez A, Bursac Z, Phillips MM, Raczynski JM. Overweight children, weight-based teasing and academic performance. Int J Pediatr Obes. 2009; 4: 274–280.

27. Gable S, Krull JL, Chang Y. Boys’ and girls’ weight status and math performance from kindergarten entry through fifth grade: a mediated analysis. Child Dev. 2012; 83(5): 1822–1839.

28. Kim TJ, Roesler NM, von dem Knesebeck O. Causation or selection - examining the relation between education and overweight/obesity in prospective observational studies: a meta-analysis. Obes Rev. 2017; 18: 660–672.

29. Finn KE, Faith MS, Seo YS. School Engagement in Relation to Body Mass Index and School Achievement in a High-School Age Sample. J Obes. 2018; 2018: 1–7.

30. Martin A, Booth JN, McGeown S, et al. Longitudinal Associations Between Childhood Obesity and Academic Achievement: Systematic Review with Focus Group Data. Curr Obes Rep. 2017; 6: 297–313.

31. Caird J, Kavanagh J, O’Mara-Eves A, et al. Does being overweight impede academic attainment? A systematic review. Health Educ. 2014; 73(5): 497–521.

32. Lindberg L, Persson M, Danielsson P, Hagman E, Marcus C. Obesity in childhood, socioeconomic status, and completion of 12 or more school years: a prospective cohort study. BMJ Open. 2021; 11: e040432.

33. Boyd A, Golding J, Macleod J, et al. Cohort Profile: the ‘children of the 90s’--the index offspring of the Avon Longitudinal Study of Parents and Children. Int J Epidemiol. 2013; 42: 111–127.

34. Fraser A, Macdonald-Wallis C, Tilling K, et al. Cohort Profile: the Avon Longitudinal Study of Parents and Children: ALSPAC mothers cohort. Int J Epidemiol. 2013; 42: 97–110.

35. Angold A, Costello EJ, Messer SC, Pickles A. Development of a short questionnaire for use in epidemiological studies of depression in children and adolescents. Int J Methods Psychiatr Res. 1995; 5: 237–249.

36. Goodman R, Ford T, Simmons H, Gatward R, Meltzer H. Using the Strengths and Difficulties Questionnaire (SDQ) to screen for child psychiatric disorders in a community sample. Br J Psychiatry. 2000; 177: 534–539.

37. Tyrrell J, Mulugeta A, Wood AR, et al. Using genetics to understand the causal influence of higher BMI on depression. Int J Epidemiol. 2019; 48(3): 834–848.

38. Rubin DB. Multiple Imputation for Nonresponse in Surveys: John Wiley & Sons; 2004.

39. Emmer C, Bosnjak M, Mata J. The association between weight stigma and mental health: A meta-analysis. Obes Rev. 2020; 21: e12935.

40. Puhl RM, Lessard LM. Weight Stigma in Youth: Prevalence, Consequences, and Considerations for Clinical Practice. Curr Obes Rep. 2020; 9: 402–411.

41. Wickersham A, Sugg HVR, Epstein S, Stewart R, Ford T, Downs J. Systematic Review and Meta-analysis: The Association Between Child and Adolescent Depression and Later Educational Attainment. J Am Acad Child Adolesc Psychiatry. 2021; 60(1): 105–118.

42. Booth JN, Tomporowski PD, Boyle JM, et al. Obesity impairs academic attainment in adolescence: findings from ALSPAC, a UK cohort. Int J Obes (Lond). 2014; 38: 1335–1342.

43. Kwong ASF, Manley D, Timpson NJ, et al. Identifying Critical Points of Trajectories of Depressive Symptoms from Childhood to Young Adulthood. J Youth Adolesc. 2019; 48: 815–827.

44. Harrison S, Davies AR, Dickson M, et al. The causal effects of health conditions and risk factors on social and socioeconomic outcomes: Mendelian randomization in UK Biobank. Int J Epidemiol. 2020; 49(5): 1661–1681.

45. Howe LD, Kanayalal R, Harrison S, et al. Effects of body mass index on relationship status, social contact and socio-economic position: Mendelian randomization and within-sibling study in UK Biobank. Int J Epidemiol. 2020; 49(4): 1173–1184.

46. Hughes A, Wade KH, Dickson M, et al. Common health conditions in childhood and adolescence, school absence, and educational attainment: Mendelian randomization study. NPJ Sci Learn. 2021; 6: 1.

47. Brumpton B, Sanderson E, Heilbron K, et al. Avoiding dynastic, assortative mating, and population stratification biases in Mendelian randomization through within-family analyses. Nat Commun. 2020; 11: 3519.

48. Blakely T, McKenzie S, Carter K. Misclassification of the mediator matters when estimating indirect effects. J Epidemiol Community Health. 2013; 67: 458–466.

49. Richiardi L, Bellocco R, Zugna D. Mediation analysis in epidemiology: methods, interpretation and bias. Int J Epidemiol. 2013; 42: 1511–1519.

50. Howe LD, Tilling K, Galobardes B, Lawlor DA. Loss to follow-up in cohort studies: bias in estimates of socioeconomic inequalities. Epidemiology. 2013; 24(1): 1–9.

