## Supplementary data for "Mediators of the association between childhood BMI and educational attainment: analysis of a UK prospective cohort study"

Affiliations

**Figure S1: Flow chart of inclusion and exclusion of ALSPAC participants into the study sample**

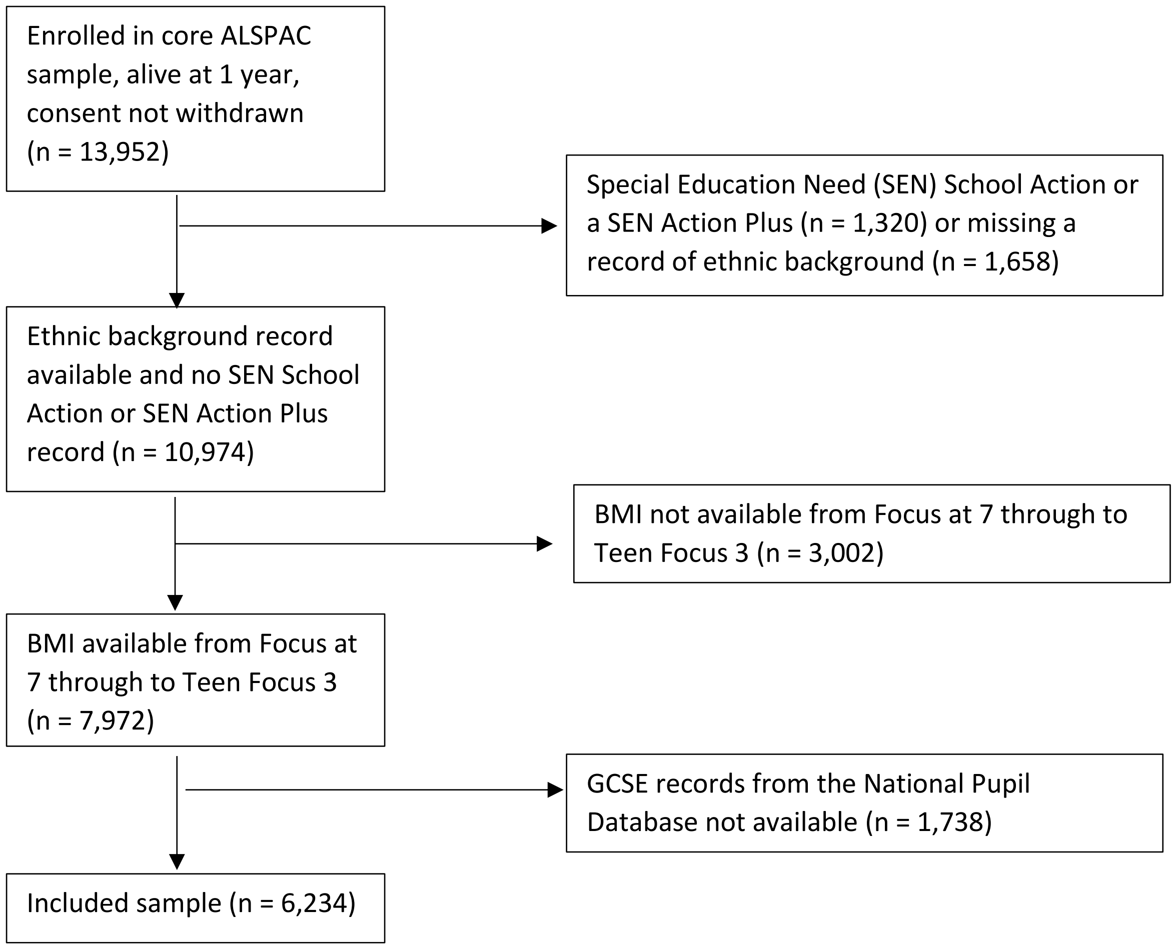

*Imputation procedure*

The predictive model contained BMI z-scores at age 11.7 years, GCSE and equivalents capped scores, depressive symptoms variables, externalising symptoms variables, school enjoyment variables, bullying variables, pressure to lose weight variables, maternal smoking in pregnancy, maternal housing tenure during pregnancy, maternal highest education qualification during pregnancy, maternal occupational social class during pregnancy, maternal age at delivery of the child, and the child's ethnicity. We also included child IQ, low birth weight (<2500 g), maternal post-natal depression as well as additional data on BMI z-scores (measures from the clinics visits from age 7.5 years to 15.5 years), depressive symptoms (Short Mood and Feelings Questionnaire scores from five points between 10 to 18.6 years), externalising symptoms (maternal reports at age six and 11 years from the strengths and difficulties questionnaire), school enjoyment (previous child reports at age 11 years), and bullying (from the clinic visits at around 8 and 10 years). Children attended a clinical assessment when they were on average 8 years old and their IQ was assessed using a short from of the Weschler Intelligence Scale for Children (WISC)-III. Imputation was carried out separately for females and males, with 20 datasets imputed.

*Table S1: Comparison of distributions for imputed data, and unimputed study sample for the female participants (n = 3,308)*

| | Percent imputed (%) | Imputed data | Observed data (study sample) |
|---|---|---|---|
| BMI z-score at 11.7 years (mean (SD)) | 21.6 | 0.32 (1.22) | 0.28 (1.19) |
| *Externalising symptoms (average age 13.2 years)* | | | |
| Teenager is generally obedient, usually does what adults request (%) | 28.6 | | |
| Certainly true | | 57.6 | 59.2 |
| Somewhat true | | 38.4 | 37.3 |
| Not true | | 4.0 | 3.4 |
| Teenager has often had temper tantrums or hot tempers (%) | 28.4 | | |
| Not true | | 55.9 | 58.0 |
| Somewhat true | | 33.8 | 33.1 |
| Certainly true | | 10.2 | 8.8 |
| Teenager often fights or bullies other children/teenagers = Certainly / Somewhat true = 1 (%) | 29.1 | 4.9 | 3.0 |
| Teenager often lies or cheats = Certainly / Somewhat true = 1 (%) | 29.2 | 15.3 | 12.3 |
| Teenager steels from home, school, elsewhere = Certainly / Somewhat true = 1 (%) | 29.0 | 4.2 | 2.2 |
| Teenager thinks things out before acting = Not true = 1 (%) | 29.1 | 10.4 | 8.9 |
| Teenager sees tasks through to end, has good attention span = Not true = 1 (%) | 28.7 | 9.7 | 8.1 |
| Teenager has been restless, overactive and can't stay still for long (%) | 28.6 | | |
| Not true | | 70.7 | 72.3 |
| Somewhat true | | 24.6 | 23.6 |
| Certainly true | | 4.7 | 4.1 |
| Teenager is constantly fidgeting or squirming (%) | 28.7 | | |
| Not true | | 81.2 | 82.4 |
| Somewhat true | | 15.4 | 14.6 |
| Certainly true | | 3.4 | 3.0 |
| Teenager is easily distracted, concentration wanders = Certainly / Somewhat true = 1(%) | 28.6 | 43.5 | 41.3 |
| *Depressive symptoms (average age 13.8 years)* | | | |
| Teenager felt miserable or unhappy in the last two weeks = Sometimes/ True = 1 (%) | 31.2 | 71.8 | 71.8 |

| | | | |
|---|---|---|---|
| Teenager didn't enjoy anything at all in the last two weeks = Sometimes/ True = 1 (%) | 31.1 | 20.6 | 19.5 |
| Teenager felt so tired he/she just sat around and did nothing in the last two weeks (%) | 31.0 | | |
| Not at all | | 48.2 | 48.4 |
| Sometimes | | 44.3 | 44.6 |
| True | | 7.5 | 7.0 |
| Teenager was very restless in the last two weeks = Sometimes/ True = 1 (%) | 31.2 | 51.7 | 51.7 |
| Teenager felt he/she was no good any more in the last two weeks (%) | 31.1 | | |
| Not at all | | 74.0 | 74.4 |
| Sometimes | | 19.0 | 19.3 |
| True | | 7.0 | 6.3 |
| Teenager cried a lot in the last two weeks = Sometimes/ True = 1 (%) | 31.1 | 32.0 | 31.7 |
| Teenager found it hard to think properly or concentrate in the last two weeks = Sometimes/ True = 1 (%) | 31.1 | 59.4 | 59.3 |
| Teenager hated him/herself in the last two weeks = Sometimes/ True = 1 (%) | 31.1 | 23.9 | 23.1 |
| Teenager was a bad person in the last two weeks = Sometimes/ True = 1 (%) | 31.1 | 21.8 | 21.3 |
| Teenager felt lonely in the last two weeks = Sometimes/ True = 1 (%) | 31.1 | 38.2 | 38.4 |
| Teenager thought nobody really loved him/her in the last two weeks = Sometimes/ True = 1 (%) | 31.1 | 23.5 | 22.4 |
| Teenager thought he/she could never be as good as other kids in the last two weeks = Sometimes/ True = 1 (%) | 31.1 | 33.1 | 32.5 |
| Teenager did everything wrong in the last two weeks = Sometimes/ True = 1 (%) | 31.1 | 25.4 | 24.3 |
| *School enjoyment (average age 14.2 years)* | | | |
| Respondent's school is a place where they really like to go each day = Strongly Disagree/ Disagree = 1 (%) | 37.1 | 36.2 | 33.4 |
| Respondent's school is a place where they get excited about the work they do (%) | 40.7 | | |
| Agree/ strongly agree | | 27.8 | 29.0 |
| Disagree | | 58.7 | 58.4 |
| Strongly disagree | | 13.6 | 12.6 |
| Respondent's school is a place where they enjoy what they do in class = Strongly Disagree/ Disagree = 1 (%) | 38.9 | 25.0 | 22.7 |
| *Bullying (average age 12.8 years)* | | | |
| Frequency someone took teenagers personal belongings (%) | 24.7 | | |

| | | | |
|---|---|---|---|
| Never (participants who responded no to preceding question) | | 78.9 | 79.8 |
| Seldom (1 to 3 times) | | 16.0 | 15.7 |
| Frequently (> 4 times)/ Very frequently (>1/week) | | 5.1 | 4.5 |
| Frequency someone threatened/ blackmailed teenager = Seldom (1 to 3 times)/ Frequently (> 4 times)/ Very frequently (>1/week) = 1 (%) | 24.6 | 9.0 | 7.9 |
| Frequency someone has tricked teenager = Seldom (1 to 3 times)/ Frequently (> 4 times)/ Very frequently (>1/week) = 1 (%) | 24.7 | 8.3 | 7.7 |
| Frequency someone has called teenager nasty names = Seldom (1 to 3 times)/ Frequently (> 4 times)/ Very frequently (>1/week) = 1 (%) | 24.8 | 40.7 | 39.6 |
| Frequency friends wouldn't hang around with teenager to upset teenager = Seldom (1 to 3 times)/ Frequently (> 4 times)/ Very frequently (>1/week) = 1 (%) | 24.8 | 14.5 | 13.6 |
| Frequency friend have tried to get teenager to do things didn't want to do = Seldom (1 to 3 times)/ Frequently (> 4 times)/ Very frequently (>1/week) = 1 (%) | 24.9 | 9.2 | 8.0 |
| Frequency friends have told lies about teenager = Seldom (1 to 3 times)/ Frequently (> 4 times)/ Very frequently (>1/week) = 1 (%) | 25.2 | 22.1 | 21.1 |
| Frequency friends have spoilt games to upset teenager: friends and peers: TF1 Seldom (1 to 3 times)/ Frequently (> 4 times)/ Very frequently (>1/week) = 1 (%) | 24.8 | 3.9 | 2.7 |
| *Pressure to lose weight (average age 13.9 years)* | | | |
| Respondent felt pressure to lose weight from friends = Yes, a lot/ Yes, quite a little/ Yes, a little = 1 (%) | 31.6 | 25.1 | 24.4 |
| Respondent felt pressure to lose weight from family = Yes, a lot/ Yes, quite a little/ Yes, a little = 1 (%) | 31.7 | 25.8 | 24.5 |
| Respondent felt pressure to lose weight from girls/boys they've gone out with = Yes, a lot/ Yes, quite a little/ Yes, a little = 1 (%) | 31.8 | 16.7 | 15.3 |
| Respondent felt pressure to lose weight from the media = Yes, a lot/ Yes, quite a little/ Yes, a little = 1(%) | 31.7 | 47.2 | 46.5 |
| Maternal smoking in pregnancy = 1 (%) | 1.4 | 18.6 | 18.5 |
| Maternal occupational social class | 14.1 | | |
| II - Managerial and technical = 1 (%) | | 30.7 | 31.2 |
| IIINM - Skilled non-manual =1 (%) | | 43.2 | 43.7 |

| | | | |
|---|---|---|---|
| IIIM - Skilled manual = 1 (%) | | 3.5 | 3.5 |
| IV - Partly skilled = 1 (%) | | 15.8 | 15.2 |
| V – Unskilled = 1 (%) | | 3.7 | 3.3 |
| Highest maternal education | 0.4 | | |
| vocational = 1 (%) | | 9.3 | 9.3 |
| O-level = 1 (%) | | 36.5 | 36.5 |
| A-level = 1 (%) | | 25.7 | 25.7 |
| Degree = 1 (%) | | 13.6 | 13.6 |
| Maternal housing tenure (Council rented/ Housing association rented) =1 (%) | 2.4 | 10.1 | 9.9 |
| Parity (≥1) = 1 (%) | 4.1 | 53.9 | 54.0 |
| Maternal age at birth (mean (SD)) | 0.0 | 28.6 (4.53) | 28.6 (4.53) |
| Capped GCSE point score (mean (SD)) | 0.0 | 353.2 (73.09) | 353.2 (73.09) |

*Table S2: Comparison of distributions for imputed data, and unimputed study sample for the male participants (n = 2,926)*

| | Percent imputed (%) | Imputed data | Observed data (study sample) |
|---|---|---|---|
| BMI z-score at 11.7 years (mean (SD)) | 22.7 | 0.41 (1.21) | 0.39 (1.18) |
| *Externalising symptoms (average age 13.2 years)* | | | |
| Teenager is generally obedient, usually does what adults request (%) | 28.0 | | |
| Certainly true | | 55.3 | 57.2 |
| Somewhat true | | 41.4 | 40.1 |
| Not true | | 3.3 | 2.8 |
| Teenager has often had temper tantrums or hot tempers (%) | 28.1 | | |
| Not true | | 59.1 | 61.4 |
| Somewhat true | | 31.8 | 30.9 |
| Certainly true | | 9.1 | 7.7 |
| Teenager often fights or bullies other children/teenagers = Certainly / Somewhat true = 1 (%) | 28.9 | 6.3 | 4.2 |
| Teenager often lies or cheats = Certainly / Somewhat true = 1 (%) | 28.5 | 19.1 | 15.7 |
| Teenager steels from home, school, elsewhere = Certainly / Somewhat true = 1 (%) | 28.6 | 4.8 | 2.3 |
| Teenager thinks things out before acting = Not true = 1 (%) | 28.7 | 13.2 | 11.4 |
| Teenager sees tasks through to end, has good attention span = Not true = 1 (%) | 27.8 | 14.3 | 12.6 |
| Teenager has been restless, overactive and can't stay still for long (%) | 28.1 | | |
| Not true | | 62.1 | 64.1 |
| Somewhat true | | 30.1 | 29.2 |
| Certainly true | | 7.8 | 6.7 |
| Teenager is constantly fidgeting or squirming (%) | 28.1 | | |
| Not true | | 71.8 | 73.4 |
| Somewhat true | | 21.9 | 21.1 |
| Certainly true | | 6.3 | 5.5 |
| Teenager is easily distracted, concentration wanders = Certainly / Somewhat true = 1(%) | 28.2 | 56.8 | 54.5 |
| *Depressive symptoms (average age 13.8 years)* | | | |
| Teenager felt miserable or unhappy in the last two weeks = Sometimes/ True = 1 (%) | 32.7 | 53.8 | 53.1 |

| | | | |
|---|---|---|---|
| Teenager didn't enjoy anything at all in the last two weeks = Sometimes/ True = 1 (%) | 32.6 | 19.6 | 18.3 |
| Teenager felt so tired he/she just sat around and did nothing in the last two weeks (%) | 32.6 | | |
| Not at all | | 48.0 | 48.5 |
| Sometimes | | 44.3 | 44.3 |
| True | | 7.7 | 7.2 |
| Teenager was very restless in the last two weeks = Sometimes/ True = 1 (%) | 32.9 | 55.2 | 54.3 |
| Teenager felt he/she was no good any more in the last two weeks (%) | 32.7 | | |
| Not at all | | 83.8 | 85.3 |
| Sometimes | | 12.8 | 12.0 |
| True | | 3.4 | 2.7 |
| Teenager cried a lot in the last two weeks = Sometimes/ True = 1 (%) | 32.6 | 7.4 | 6.4 |
| Teenager found it hard to think properly or concentrate in the last two weeks = Sometimes/ True = 1 (%) | 32.7 | 53.6 | 53.2 |
| Teenager hated him/herself in the last two weeks = Sometimes/ True = 1 (%) | 32.7 | 11.9 | 10.4 |
| Teenager was a bad person in the last two weeks = Sometimes/ True = 1 (%) | 32.8 | 16.9 | 15.7 |
| Teenager felt lonely in the last two weeks = Sometimes/ True = 1 (%) | 32.7 | 23.1 | 22.5 |
| Teenager thought nobody really loved him/her in the last two weeks = Sometimes/ True = 1 (%) | 32.7 | 10.5 | 9.6 |
| Teenager thought he/she could never be as good as other kids in the last two weeks = Sometimes/ True = 1 (%) | 32.7 | 19.6 | 18.6 |
| Teenager did everything wrong in the last two weeks = Sometimes/ True = 1 (%) | 32.7 | 13.2 | 12.1 |
| *School enjoyment (average age 14.2 years)* | | | |
| Respondent's school is a place where they really like to go each day = Strongly Disagree/ Disagree = 1 (%) | 48.2 | 37.6 | 35.4 |
| Respondent's school is a place where they get excited about the work they do (%) | 50.3 | | |
| Agree/ strongly agree | | 29.8 | 29.6 |
| Disagree | | 57.4 | 58.1 |
| Strongly disagree | | 12.9 | 12.3 |
| Respondent's school is a place where they enjoy what they do in class = Strongly Disagree/ Disagree = 1 (%) | 49.0 | 26.3 | 23.0 |
| *Bullying (average age 12.8 years)* | | | |
| Frequency someone took teenagers personal belongings (%) | 26.1 | | |

| | | | |
|---|---|---|---|
| Never (participants who responded no to preceding question) | | 75.6 | 76.0 |
| Seldom (1 to 3 times) | | 17.4 | 17.3 |
| Frequently (> 4 times)/ Very frequently (>1/week) | | 6.9 | 6.6 |
| Frequency someone threatened/ blackmailed teenager = Seldom (1 to 3 times)/ Frequently (> 4 times)/ Very frequently (>1/week) = 1 (%) | 26.0 | 11.1 | 9.8 |
| Frequency someone has tricked teenager = Seldom (1 to 3 times)/ Frequently (> 4 times)/ Very frequently (>1/week) = 1 (%) | 26.0 | 8.3 | 7.5 |
| Frequency someone has called teenager nasty names = Seldom (1 to 3 times)/ Frequently (> 4 times)/ Very frequently (>1/week) = 1 (%) | 26.2 | 33.9 | 33.1 |
| Frequency friends wouldn't hang around with teenager to upset teenager = Seldom (1 to 3 times)/ Frequently (> 4 times)/ Very frequently (>1/week) = 1 (%) | 26.3 | 7.7 | 6.6 |
| Frequency friend have tried to get teenager to do things didn't want to do = Seldom (1 to 3 times)/ Frequently (> 4 times)/ Very frequently (>1/week) = 1 (%) | 26.3 | 8.4 | 7.2 |
| Frequency friends have told lies about teenager = Seldom (1 to 3 times)/ Frequently (> 4 times)/ Very frequently (>1/week) = 1 (%) | 26.6 | 12.8 | 12.2 |
| Frequency friends have spoilt games to upset teenager: friends and peers: TF1 Seldom (1 to 3 times)/ Frequently (> 4 times)/ Very frequently (>1/week) = 1 (%) | 26.2 | 7.8 | 7.0 |
| *Pressure to lose weight (average age 13.9 years)* | | | |
| Respondent felt pressure to lose weight from friends = Yes, a lot/ Yes, quite a little/ Yes, a little = 1 (%) | 43.0 | 17.1 | 15.6 |
| Respondent felt pressure to lose weight from family = Yes, a lot/ Yes, quite a little/ Yes, a little = 1 (%) | 42.9 | 16.6 | 15.0 |
| Respondent felt pressure to lose weight from girls/boys they've gone out with = Yes, a lot/ Yes, quite a little/ Yes, a little = 1 (%) | 42.9 | 9.3 | 7.7 |
| Respondent felt pressure to lose weight from the media = Yes, a lot/ Yes, quite a little/ Yes, a little = 1(%) | 42.9 | 13.8 | 12.1 |
| Maternal smoking in pregnancy = 1 (%) | 1.4 | 18.6 | 18.5 |
| Maternal social class | 11.8 | | |
| Maternal social class (II - Managerial and technical) = 1 (%) | | 30.7 | 31.1 |

| | | | |
|---|---|---|---|
| Maternal social class (IIINM - Skilled non-manual) =1 (%) | | 44.3 | 44.8 |
| Maternal social class (IIIM - Skilled manual) = 1 (%) | | 3.8 | 3.8 |
| Maternal social class (IV - Partly skilled) = 1 (%) | | 14.5 | 14.0 |
| Maternal social class (V – Unskilled) = 1 (%) | | 2.9 | 2.6 |
| Highest maternal education | 0.1 | | |
| Highest maternal education (vocational) = 1 (%) | | 9.1 | 9.1 |
| Highest maternal education (O-level) = 1 (%) | | 36.9 | 36.9 |
| Highest maternal education (A-level) = 1 (%) | | 26.0 | 26.0 |
| Highest maternal education (Degree) = 1 (%) | | 14.0 | 14.0 |
| Maternal housing tenure (Council rented/ Housing association rented) =1 (%) | 2.3 | 9.0 | 8.9 |
| Parity (≥1) = 1 (%) | 3.7 | 53.5 | 53.4 |
| Maternal age at birth (mean (SD)) | 0.0 | 29.0 (4.59) | 29.0 (4.59) |
| Capped GCSE point score (mean (SD)) | 0.0 | 337.9 (79.02) | 337.9 (79.02) |

*Table S3: Model fit statistics for the female (n = 3,308) and male (n = 2,926) sample from the measurement part of the structural equation models (SEM)*

| | **Females measurement model** | | | **Males measurement model** | | |
|---|---|---|---|---|---|---|
| | CFI median (lower quartile - upper quartile) | TLI median (lower quartile - upper quartile) | RMSEA median (lower quartile - upper quartile) | CFI median (lower quartile - upper quartile) | TLI median (lower quartile - upper quartile) | RMSEA median (lower quartile - upper quartile) |
| Depressive symptoms | 0.993 (0.992 - 0.993) | 0.992 (0.991 - 0.992) | 0.040 (0.039 - 0.041) | 0.989 (0.988 - 0.990) | 0.987 (0.986 - 0.988) | 0.037 (0.036 - 0.038) |
| School enjoyment | NA | NA | NA | NA | NA | NA |
| Pressure to lose weight | NA | NA | NA | NA | NA | NA |
| Externalising symptoms | 0.931 (0.927 - 0.934) | 0.911 (0.906 - 0.915) | 0.084 (0.080 - 0.086) | 0.939 (0.935 - 0.942) | 0.922 (0.916 - 0.926) | 0.084 (0.082 - 0.086) |
| Bullying | 0.996 (0.995 - 0.997) | 0.994 (0.993 - 0.995) | 0.020 (0.018 - 0.023) | 0.998 (0.997 - 0.999) | 0.997 (0.996 - 0.999) | 0.013 (0.010 - 0.016) |

NA is shown where it is not possible to assess model fit due to a small number of degrees of freedom; there are only 3 variables informing the latent variable for school enjoyment, and only 4 variables informing the latent variable for pressure to lose weight

*Table S4: Unstandardized factor loadings for externalising symptoms, depressive symptoms, school enjoyment, bullying/peer victimisation, and pressure to lose weight from the female (n = 3,308) and male (n = 2,926) sample from measurement part of the structural equation models (SEM)*

| | **Females measurement model** | **Males measurement model** |
|---|---|---|
| | median (lower quartile - upper quartile) | median (lower quartile - upper quartile) |
| *Externalising symptoms* | | |
| Teenager is generally obedient, usually does what adults request | 1.00 (1.00 - 1.00) | 1.00 (1.00 - 1.00) |
| Teenager has often had temper tantrums or hot tempers | 1.07 (1.03 - 1.08) | 1.11 (1.08 - 1.13) |
| Teenager often fights or bullies other children/teenagers | 1.17 (1.14 - 1.23) | 1.10 (1.05 - 1.12) |
| Teenager often lies or cheats | 1.32 (1.28 - 1.36) | 1.25 (1.19 - 1.27) |
| Teenager steels from home, school, elsewhere | 1.05 (0.99 - 1.12) | 0.95 (0.87 - 1.02) |
| Teenager thinks things out before acting | 1.16 (1.13 - 1.19) | 1.28 (1.25 - 1.31) |
| Teenager sees tasks through to end, has good attention span | 1.17 (1.15 - 1.21) | 1.21 (1.17 - 1.24) |
| Teenager has been restless, overactive and can't stay still for long | 1.32 (1.29 - 1.34) | 1.39 (1.35 - 1.41) |
| Teenager is constantly fidgeting or squirming | 1.37 (1.34 - 1.39) | 1.42 (1.37 - 1.45) |
| Teenager is easily distracted, concentration wanders | 1.05 (1.02 - 1.07) | 1.17 (1.12 - 1.20) |
| *Depressive symptoms* | | |
| Teenager felt miserable or unhappy in the last two weeks | 1.00 (1.00 - 1.00) | 1.00 (1.00 - 1.00) |
| Teenager didn't enjoy anything at all in the last two weeks | 0.78 (0.77 - 0.80) | 0.86 (0.83 - 0.87) |

| | | |
|---|---|---|
| Teenager felt so tired he/she just sat around and did nothing in the last two weeks | 0.60 (0.58 - 0.62) | 0.69 (0.66 - 0.72) |
| Teenager was very restless in the last two weeks | 0.66 (0.64 - 0.68) | 0.64 (0.63 - 0.66) |
| Teenager felt he/she was no good any more in the last two weeks | 1.20 (1.19 - 1.22) | 1.26 (1.25 - 1.29) |
| Teenager cried a lot in the last two weeks | 0.97 (0.95 - 0.97) | 1.05 (1.02 - 1.07) |
| Teenager found it hard to think properly or concentrate in the last two weeks | 0.79 (0.77 - 0.80) | 0.93 (0.91 - 0.96) |
| Teenager hated him/herself in the last two weeks | 1.19 (1.18 - 1.21) | 1.28 (1.26 - 1.30) |
| Teenager was a bad person in the last two weeks | 0.96 (0.95 - 0.97) | 0.97 (0.94 - 1.00) |
| Teenager felt lonely in the last two weeks | 1.06 (1.04 - 1.07) | 1.19 (1.17 - 1.20) |
| Teenager thought nobody really loved him/her in the last two weeks | 1.11 (1.09 - 1.12) | 1.27 (1.23 - 1.30) |
| Teenager thought he/she could never be as good as other kids in the last two weeks | 1.06 (1.05 - 1.07) | 1.15 (1.13 - 1.17) |
| Teenager did everything wrong in the last two weeks | 1.10 (1.08 - 1.11) | 1.18 (1.15 - 1.22) |
| *School enjoyment* | | |
| Respondent's school is a place where they really like to go each day (%) | 1.00 (1.00 - 1.00) | 1.00 (1.00 - 1.00) |
| Respondent's school is a place where they get excited about the work they do (%) | 1.00 (0.98 - 1.02) | 0.93 (0.91 - 0.94) |
| Respondent's school is a place where they enjoy what they do in class (%) | 1.20 (1.19 - 1.22) | 1.15 (1.13 - 1.18) |
| *Bullying* | | |

| | | |
|---|---|---|
| Frequency someone took teenagers personal belongings: friends and peers | 1.00 (1.00 - 1.00) | 1.00 (1.00 - 1.00) |
| Frequency someone threatened/ blackmailed teenager: friends and peers | 1.12 (1.08 - 1.15) | 1.21 (1.19 - 1.23) |
| Frequency someone has tricked teenager: friends and peers | 1.16 (1.12 - 1.17) | 1.22 (1.19 - 1.26) |
| Frequency someone has called teenager nasty names: friends and peers | 1.09 (1.09 - 1.12) | 1.21 (1.19 - 1.23) |
| Frequency friends wouldn't hang around with teenager to upset teenager: friends and peers | 1.20 (1.18 - 1.22) | 1.31 (1.28 - 1.32) |
| Frequency friend have tried to get teenager to do things didn't want to do | 1.18 (1.15 - 1.20) | 1.22 (1.20 - 1.24) |
| Frequency friends have told lies about teenager: friends and peers | 1.31 (1.29 - 1.34) | 1.26 (1.23 - 1.28) |
| Frequency friends have spoilt games to upset teenager: friends and peers | 0.86 (0.80 - 0.90) | 1.03 (0.99 - 1.06) |
| *Pressure to lose weight* | | |
| Respondent felt pressure to lose weight from friends | 1.00 (1.00 - 1.00) | 1.00 (1.00 - 1.00) |
| Respondent felt pressure to lose weight from family | 0.78 (0.77 - 0.80) | 0.86 (0.85 - 0.87) |
| Respondent felt pressure to lose weight from girls/boys they've gone out with | 0.92 (0.91 - 0.94) | 0.80 (0.78 - 0.82) |
| Respondent felt pressure to lose weight from the media | 0.90 (0.89 - 0.92) | 0.80 (0.79 - 0.82) |

*Table S5: Pooled total effect, remaining direct effect, indirect effect, and proportion mediated restricting to those with a white British ethnic background for females (n = 3,177) and males (n = 2,803) from the structural equation models (SEM)*

| | Total effect | | | Remaining direct effect | | | Indirect effect | | | Proportion mediated |
|---|---|---|---|---|---|---|---|---|---|---|
| | β[1] | 95% CI β | p-value | β[1] | 95% CI | p-value | β[1] | 95% CI | p-value | |
| **Females** | | | | | | | | | | |
| | -3.34 | ( -5.29 to -1.39 ) | 7.85e-04 | | | | | | | |
| Depressive Symptoms | | | | -2.96 | ( -4.92 to -1.01 ) | 2.94e-03 | -0.38 | ( -0.68 to -0.07 ) | 1.60e-02 | 0.11 |
| School enjoyment | | | | -3.41 | ( -5.31 to -1.50 ) | 4.55e-04 | 0.07 | ( -0.77 to 0.90 ) | 8.77e-01 | NA |
| Pressure to lose weight | | | | -3.70 | ( -6.29 to -1.12 ) | 5.03e-03 | 0.36 | ( -1.34 to 2.06 ) | 6.77e-01 | NA |
| Externalising symptoms | | | | -2.52 | ( -4.40 to -0.63 ) | 8.75e-03 | -0.83 | ( -1.74 to 0.09 ) | 7.58e-02 | 0.25 |
| Bullying | | | | -3.26 | ( -5.22 to -1.31 ) | 1.08e-03 | -0.08 | ( -0.28 to 0.13 ) | 4.64e-01 | 0.02 |
| **Males** | | | | | | | | | | |
| | -2.33 | ( -4.53 to -0.12 ) | 3.84e-02 | | | | | | | |
| Depressive Symptoms | | | | -2.02 | ( -4.20 to 0.17 ) | 7.05e-02 | -0.31 | ( -0.88 to 0.25 ) | 2.78e-01 | 0.13 |
| School enjoyment | | | | -2.35 | ( -4.57 to -0.13 ) | 3.79e-02 | 0.02 | ( -0.52 to 0.56 ) | 9.36e-01 | NA |
| Pressure to lose weight | | | | -0.70 | ( -4.29 to 2.89 ) | 7.01e-01 | -1.63 | ( -4.47 to 1.22 ) | 2.63e-01 | 0.70 |
| Externalising symptoms | | | | -1.94 | ( -4.06 to 0.17 ) | 7.16e-02 | -0.39 | ( -1.50 to 0.72 ) | 4.95e-01 | 0.17 |
| Bullying | | | | -1.94 | ( -4.16 to 0.27 ) | 8.52e-02 | -0.39 | ( -0.80 to 0.03 ) | 6.57e-02 | 0.17 |

[1] unstandardized

*SEM models were adjusted for maternal age at pregnancy (years), maternal smoking in pregnancy, housing tenure, highest maternal education qualification, maternal social class, and parity.*
